# Rt2: computing and visualising COVID-19 epidemics temporal reproduction number

**DOI:** 10.1101/2020.12.05.20244376

**Authors:** Bastien Reyné, Gonché Danesh, Samuel Alizon, Mircea T. Sofonea

**Affiliations:** MIVEGEC, CNRS, IRD, Université de Montpellier, 911 avenue Agropolis, 34394 Montpellier, France; MIVEGEC, Université de Montpellier, CNRS, IRD, 911 avenue Agropolis, 34394 Montpellier, France

## Abstract

Analysing the spread of COVID-19 epidemics in a timely manner is essential for public health authorities. However, raw numbers may be misleading because of spatial and temporal variations. We introduce Rt2, an R-program with a shiny interface, which uses incidence data, i.e. number of new cases per day, to compute variations in the temporal reproduction number (ℛ_*t*_), which corresponds to the average number of secondary infections caused by an infected person. This number is computed with the **R0** package, which better captures past variations, and the **EpiEstim** package, which provides a more accurate estimate of current values. ℛ_*t*_ can be computed in different countries using either the daily number of new cases or of deaths. For France, these numbers can also be computed at the regional and departmental level using also daily numbers of hospital and ICU admissions. Finally, in addition to ℛ_*t*_, we represent the incidence using a one-week sliding window to buffer daily variations. Overall, Rt2 provides an accurate and timely overview of the state and speed of spread of COVID-19 epidemics at different scales, using different metrics.

## Context

Monitoring the state and speed of spread of COVID-19 epidemics at the national and regional levels is crucial to implement non-pharmaceutical interventions (Flaxman et al., 2020). Every day, public health agencies communicate key figures to monitor the epidemic, especially incidences, which correspond to the number of new cases detected. These incidences are typically related to four variables, which are PCR-based detection, deaths, hospitalisations, and ICU admissions. The statistical analysis of time variations in these time series can inform us about epidemiological dynamics.

The purpose of the Rt2 application is to visualize the trends present in COVID-19 data through the temporal reproduction number, noted ℛ(*t*) (Wallinga and Lipsitch, 2007). This is done at different levels (country and, for France, region, and department) using different types of data (incidence in number of cases detected, in number of deaths and, for France, in number of hospitalisations, and ICU admissions).

## Reproduction numbers

### Overview

One of the key parameters in an epidemic is the reproduction number, ℛ, which reads as the average number of people infected by a contagious person during his or her infection. At the beginning of an epidemic, when the whole population is susceptible (*i*.*e*. not immune) and no interventions are implemented, this number has a particular value noted ℛ_0_ and called the basic reproduction number. During the course of the epidemic, when the proportion of immunized people becomes large enough to slow the transmission of the virus (by an effect comparable to a dilution of the individuals still susceptible), this number is called the effective, or temporal reproduction number, and denoted ℛ(*t*).

Intuitively, if ℛ(*t*) > 1, then one person is infecting more than one person on average and the epidemic is growing, as shown in textbooks such as the one by Anderson and May (1991). As the COVID-19 epidemic spreads, ℛ(*t*) decreases as an increasing proportion of the population becomes immune. When the group immunity threshold is exceeded, ℛ(*t*) falls below the threshold of 1, an epidemic peak is reached, and the epidemic declines. Public health control measures can also decrease ℛ(*t*). Therefore, the epidemic peak can be reached before the threshold for herd immunity is.

At a given point in time *t*, knowing the value of ℛ(*t*) is therefore essential to determine the status of the epidemic.

### Formal definitions

In practice, as discussed by Wallinga and Lipsitch (2007), we need two pieces of information to calculate the reproduction number:

1. the growth rate of the epidemic at a given time, *r*(*t*), which can be easily calculated from incidence time series (daily number of new cases detected, new hospitalizations, new deaths…),
2. the time during which a person is contagious, *D*.

A well-known and intuitive relationship between all these terms is as follows:

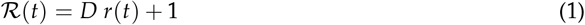

However, this apparent simplicity masks significant problems.

First, even if we assume a Markovian setting, *i*.*e*. in the absence of memory discussed for COVID-19 epidemics by Sofonea et al. (2020), this equation remains an approximation: the ℛ(*t*) from equation 2 is underestimated by a quantity ℛ_0_ *p* (*t*), where *p* (*t*) refers to the proportion of cases at time *t*, a quantity that is not always negligible, especially near the epidemic peak.

Second, the duration of contagiousness *D* is difficult to capture. It is tempting to use a fixed duration but this implicitly assumes that individuals become contagious immediately after infection and lose contagiousness at a constant rate (Markovian regime); two conditions that are rarely satisfied. On the one hand, there is often a latency period during which the individual is not infectious (in the case of COVID-19, this latency is less than the incubation period, hence the difficulty of measurement, as shown by He et al. (2020)). This imposes a positive corrective term to the estimated growth rate. On the other hand, the loss of contagiousness seems to show a non-Markovian pattern (He et al., 2020).

To visualize the neither exponential nor uniform aspect of the distribution of contagiousness over time, we show below the COVID-19 serial interval compiled from 28 infector-infectee pairs by Nishiura et al. (2020) in Figure 1. Clearly, the median (in red) and even 95% confidence interval (between the blue bounds) are both not very representative of the totality of the values.

**Figure 1:**
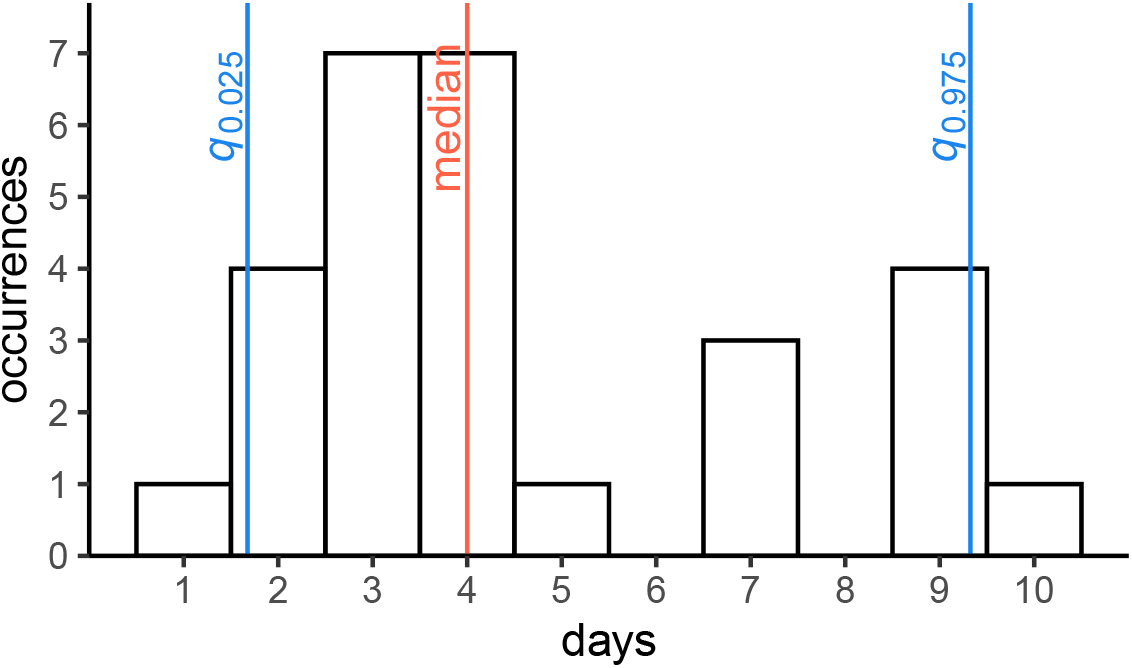
Serial interval for COVID-19 epidemics computed by Nishiura et al. (2020). The blue lines show the 95% confidence interval and the red line shows the median.

To circumvent the contagiousness duration problem and its modelling in a classical SIR model, while relying on the empirical distribution, an elegant solution is provided by the Euler-Lotka equation. As detailed by Wallinga and Lipsitch (2007), provided that the epidemic growth is exponential, we can write

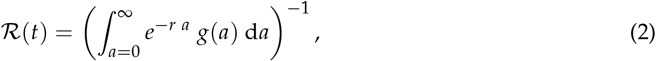

where *a* is the ‘age’ of an infection, *g*(*a*) is the distribution of contagiousness durations, and *r* is the growth rate, assumed to be constant over the time interval. In practice, *g*(*a*) is approximated by the serial interval, noted here as *w*(*a*), described above and which corresponds to the time between the onset of symptoms in an infection pair.

Outside the exponential regime and, in particular, in the vicinity of an epidemic peak, the continuous formulation in equation 2 does not apply. However, we can use a general formula introduced by Wallinga and Lipsitch (2007) that directly involves incidence data ((*y*_1_, …, *y*_*n*_), where *y*_*k*_ represents the number of new cases detected on day *k*):

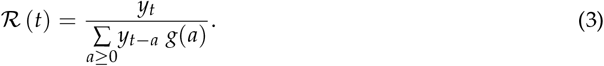

The latter expression also has the advantage to work in discrete time, which is more appropriate for incidence data.

### Implementation

Building on existing R-packages, we developed a shiny interface to show the temporal reproduction number and the incidence for a variety of data at different scales. We briefly present the data and the packages used, as well as the shiny implementation itself.

### Data processing

Data is collected from two repositories: that from Max Roser and Hasell (2020), which itself mainly uses the data from the European Centre for Disease Prevention and Control (ECDC), and, for France, the national public health agency Santé Publique France. Both sources provide a daily-updated CSV file.

Since the packages used to compute an estimation of the temporal reproduction number require a two-columns data frame as an input (date and incidence data), raw data were pre-processed using **dplyr** and **tidyr** packages. More specifically, we created a function returning the desired data frame given the type of data and the location wanted.

It also might worth to note the incidence data were smoothed out using a 7-days rolling average, in order to compensate the incidence reporting delays happening on week-ends. We used the rollmean function from the **zoo** package.

### R0

The **R0** R-package developed by Obadia et al. (2012) implements maximum likelihood estimation methods for the basic reproduction number ℛ_0_ using the approach from White and Pagano (2008).

It can also compute temporal reproduction numbers ℛ(*t*) using a procedure introduced by Wallinga and Teunis (2004), which works in the following way. We assume an epidemic curve is a time series of incidences (*y*_1_, …, *y*_*n*_), where *y*_*k*_ represents the number of new cases detected on day *k*. Note that in the original model, detection corresponds to the onset of symptoms.

This time series is the result of a set of transmission events between all cases detected since the beginning of the epidemic. If the proportion of undetected cases remains constant over the time window studied, the estimate is not biased; otherwise, pre-processing of the data is necessary, as in White et al. (2009). The structure of this set of events is seen as a tree (more precisely a connex acyclic oriented graph) whose probability of underlying data (*i*.*e*. the likelihood) is decomposed into infector/infected pairs (transmissions are assumed to be independent).

If *p*_*i,j*_ denotes the probability that an individual I_*d*_ detected on day *t*_*d*_ is the infector of an individual I_*i*_ detected on the day *t*_*i*_, then, by definition of the serial interval the distribution of which is characterized thereafter (*w*_*k*_)_*k*_ ≥ 0, we have

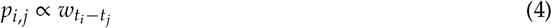

Therefore, the relative likelihood *l*_*i,j*_ of the pair (*i, j*) to be a transmission pair is given by the expression

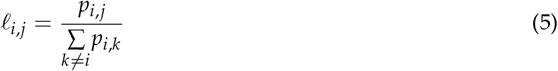

The individual reproduction number of case *j* (denoted *R*_*j*_) is therefore the sum of all cases *i* she/he may have infected, weighted by the relative likelihood of each pair:

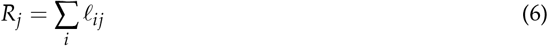

The temporal reproduction number is obtained by averaging the individual reproduction number over all cases detected on the same day *j*, where *t*_*j*_ = *t*:

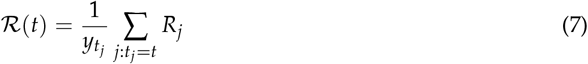

Note that **R0** allows the user to enter raw values for the serial interval (which we implement in one of the options of Rt2).

### EpiEstim

The **EpiEstim** software package, developed by Cori et al. (2013) and updated by Thompson et al. (2019), is motivated by the fact that, in situations where the studied epidemic is still ongoing, the total number of infections caused by the last detected cases is not yet known. This issue is particularly acute when it comes to evaluating the effectiveness of control measures rapidly (i.e. within few days).

In the approach used by Wallinga and Teunis (2004) and Obadia et al. (2012) (in the **R0** package), the reproduction number of cases (or of cohorts), is retrospective: its calculation is based on the number of secondary cases actually caused by a cohort of detected infectors from the date on which they were detected.

Conversely, the method used by Cori et al. (2013) and Thompson et al. (2019) in the **EpiEstim** package computes the instantaneous reproduction number in a prospective manner: its calculation is based on the potential number of secondary infections that a cohort of cases could have caused if the conditions of transmissibility had remained the same as at the time of their detection.

Formally **EpiEstim** maximizes the likelihood of incidence data (viewed as a Poisson count) observed over a time window in which the reproduction number is assumed to be constant. By noting *y*_*k*_ and 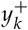 respectively the numbers of new local and total (i.e. including imported) cases, and *w*_*s*_ the probability corresponding to a serial interval *s*, the temporal (instantaneous) reproduction number for the interval [*t*; *t* − *τ*] satisfies

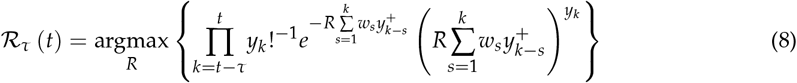

Quoting Cori et al. (2013), this distinction is equivalent to that between the (retrospective) life expectancy of a cohort of individuals, calculated once they have all died, and the (prospective) life expectancy of the same cohort, estimated under the assumption that mortality will remain the same as that known at birth. Note that the additional calculations required by the **EpiEstim** software package to make inferences tend to increase the computation time.

### Interface

We used the **shiny** package in order to present the results through an interactive web app allowing users to choose key parameters. Unfortunately, the **EpiEstim** package proved to be too slow for such an interactive setting. To overcome this issue, we first generate a CSV file that contains every single parameter combination and can be read by the app. The CSV file is routinely updated on a day-to-day basis.

### Visualisation

The visualisation was performed using the **dygraphs** library, integrated to the shiny app through the **htmltools** package. The **dygraphs** package allows for interactions, mainly zooming-in on the time axis (*x*-axis) and displaying the value of any mouse-selected day.

Figure 2 shows a typical screenshot of the visual interface.

**Figure 2:**
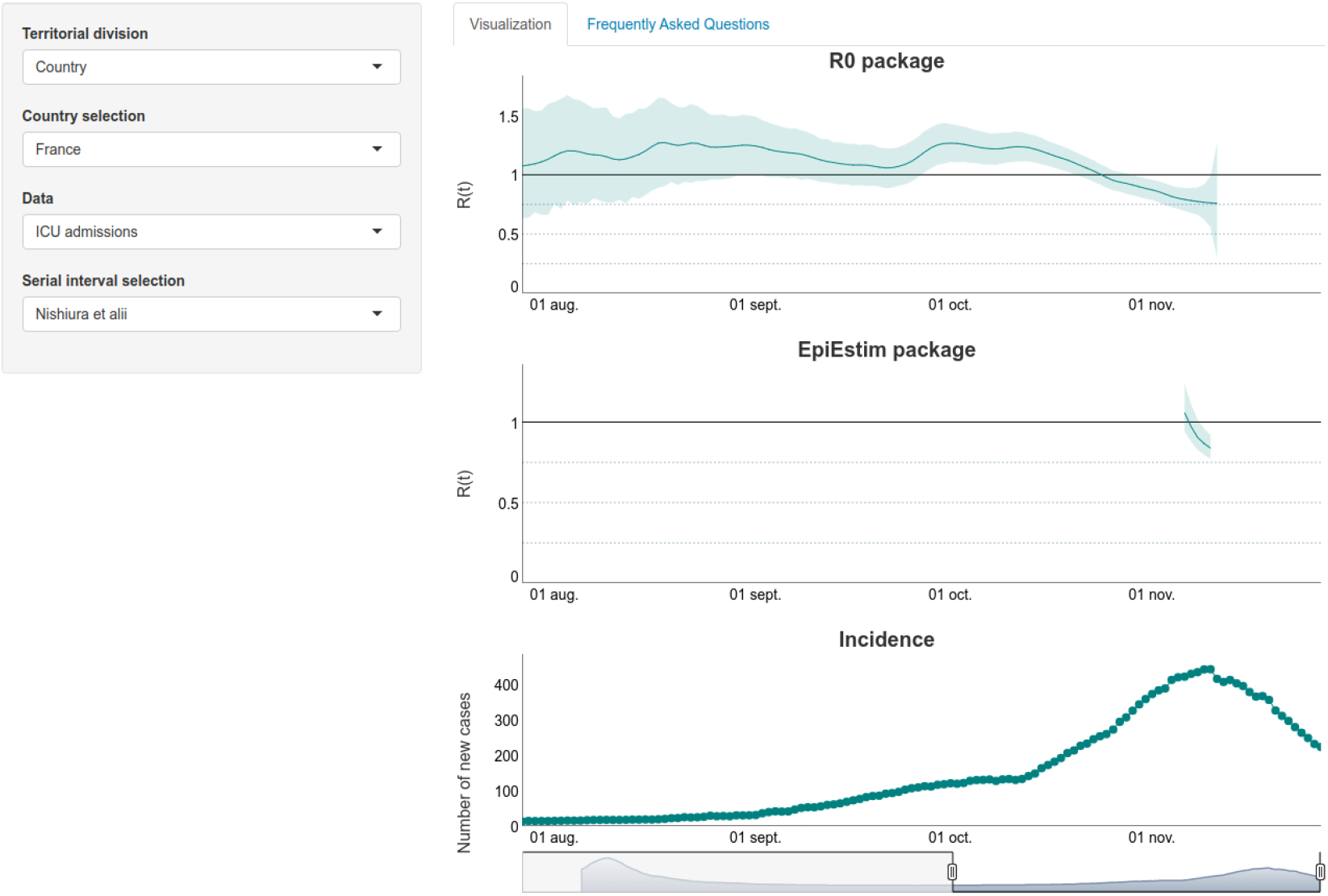
Screenshot of the Rt2 for ICU admissions incidence data from France.

### Limitations and interpretations

There are a series of limitations inherent to the estimation of the temporal reproduction number (ℛ*t*), which we attempted to address in the development of Rt2 to limit misinterpretation risks.

### Time delays

While the reproduction number quantifies a potential for the spread of the epidemic at a given date, its estimate is based on data that reflect the state of the epidemic several days earlier. Indeed, from the moment a person is infected, it takes a few days before the virus can be detected, and then usually another week before a potential hospitalization, with a possible admission in an intensive care unit (ICU) after another delay. Death can occur several days or weeks after ICU admission. In addition, time must be allowed for the case/admission/death to be identified and reported.

To account for this, and based on estimations of the COVID-19 epidemics in France (Sofonea et al., 2020), we impose a shift of 10, 12, 14, and 28 days between ℛ(*t*) and the case, hospitalisation, ICU admission, and death daily incidence data respectively.

### Sampling effort variations

While it is not necessary for all cases to be recorded for the methods in **R0** and **EpiEstim** to work properly, it is necessary for the sampling rate to be as constant as possible over time. Indeed, an intensification of screening effort mechanically amplifies the number of cases detected, therefore mimicking a growing epidemic. In France, case incidence in can be particularly misleading because screening intensity was initially very low (limited to severe cases), but increased a lot during the epidemic. Furthermore, there are known weekly variations with lower incidences during week-ends for instance.

To limit variations in sampling efforts, we use a 7-days moving average in our calculations and when plotting the incidence data.

### Serial interval

As explained above, a key input for estimating the reproduction number is the number of days during which a person is contagious. In practice, this ‘generation time’ is estimated by tracking contacts (*i*.*e*. infector-infectee pairs) and counting the number of days between the dates of onset of symptoms in the infecting and infected individuals respectively. This approximation via the symptom onset dates corresponds to the the serial interval.

Data for this serial interval in France and in Europe was initially (and still largely is) very limited. To account for this, our application allows the user to choose between several data sets for the serial interval in order to better understand the effect of this parameter on the results shown in Figure 3. Note that one of the distributions available corresponds to the raw data from Nishiura et al. (2020).

**Figure 3:**
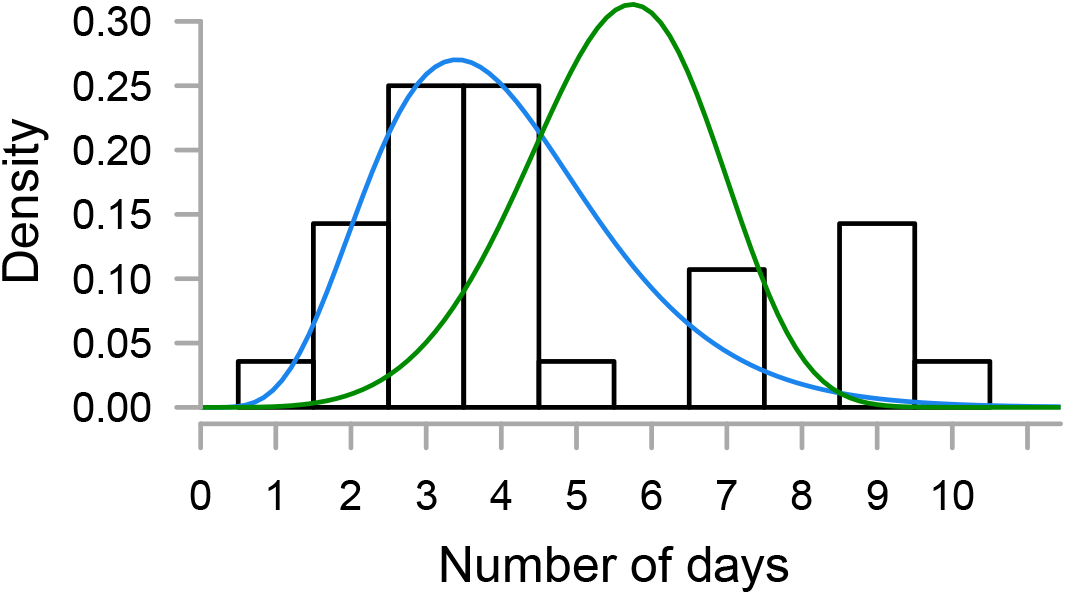
Serial intervals available in the Rt2 package. The blue curve is a Gamma distribution, Gamma(6.5, 0.62), the green curve is a Weibull distribution, Weibull(5, 6), and the histogram shows the raw data from Nishiura et al. (2020).

### Summary

We developed an R-program with a shiny interface, which shows variation in incidence data and in the temporal reproduction numbers (ℛ_*t*_). The originality is that we use different methods to compute the reproduction number, and allow the use to select incidence data from various geographical locations (country, region, department) or from different types (infections, deaths, hospitalisations, ICU admissions). Furthermore, the Rt2 interface is built to minimise the risk of misinterpretation related to variations in sampling effort, delays between incidence data and epidemic state, and serial interval. This tool can directly be used by public health authorities or the general public.

## Data Availability

All the data used in this study are publicly available.

https://bioinfo-shiny.ird.fr/Rt2-en/

## Acknowledgements

We are very grateful to the the itrop HPC (South Green Platform) of IRD Montpellier for hosting this application (more details on https://bioinfo.ird.fr/).

The ETE modeling team is composed of Samuel Alizon, Thomas Bénéteau, Marc Choisy, Gonché Danesh, Ramsès Djidjou-Demasse, Baptiste Elie, Yannis Michalakis, Bastien Reyné, Quentin Richard, Christian Selinger, Mircea T. Sofonea.

This work was partly supported by the Occitanie region and the ANR (PHYEPI project). GD is funded by the Fondation pour la Recherche Médicale (FRM grant number ECO20170637560).

